# Incidence of COVID-19 and Connections with Air Pollution Exposure: Evidence from the Netherlands

**DOI:** 10.1101/2020.04.27.20081562

**Authors:** Bo Pieter Johannes Andrée

## Abstract

The fast spread of severe acute respiratory syndrome coronavirus 2 has resulted in the emergence of several hot-spots around the world. Several of these are located in areas associated with high levels of air pollution. This study investigates the relationship between exposure to particulate matter and COVID-19 incidence in 355 municipalities in the Netherlands. The results show that atmospheric particulate matter with diameter less than 2.5 is a highly significant predictor of the number of confirmed COVID-19 cases and related hospital admissions. The estimates suggest that expected COVID-19 cases increase by nearly 100 percent when pollution concentrations increase by 20 percent. The association between air pollution and case incidence is robust in the presence of data on health-related preconditions, proxies for symptom severity, and demographic control variables. The results are obtained with ground-measurements and satellite-derived measures of atmospheric particulate matter as well as COVID-19 data from alternative dates. The findings call for further investigation into the association between air pollution and SARS-CoV-2 infection risk. If particulate matter plays a significant role in COVID-19 incidence, it has strong implications for the mitigation strategies required to prevent spreading.

**Highlights:** *Background:* Research on viral respiratory infections has found that infection risks increase following exposure to high concentrations of particulate matter. Several hot-spots of Severe Acute Respiratory Syndrome Coronavirus 2 infections are in areas associated with high levels of air pollution.

*Approach:* This study investigates the relationship between exposure to particulate matter and COVID-19 incidence in 355 municipalities in the Netherlands using data on confirmed cases and hospital admissions coded by residence, along with local PM_2.5_, PM_10_, population density, demographics and health-related pre-conditions. The analysis utilizes different regression specifications that allow for spatial dependence, nonlinearity, alternative error distributions and outlier treatment.

*Results:* PM_2.5_ is a highly significant predictor of the number of confirmed COVID-19 cases and related hospital admissions. Taking the WHO guideline of 10mcg/m3 as a baseline, the estimates suggest that expected COVID-19 cases increase by nearly 100% when pollution concentrations increase by 20%.

*Conclusion:* The findings call for further investigation into the association between air pollution on SARS-CoV-2 infection risk. If particulate matter plays a significant role in the incidence of COVID-19 disease, it has strong implications for the mitigation strategies required to prevent spreading, particularly in areas that have high levels of pollution.

## 1. Introduction

In 2019, confirmed infections with a new novel human coronavirus (SARS-CoV-2) emerged in Wuhan, in the Hubei Province in China. The virus rapidly spread to other parts of China and by early 2020 it had emerged in many other countries around the world. The World Health Organization (WHO) declared a global pandemic on March 11 2020, as confirmed cases topped 118,000 in more than 110 countries and territories around the world with sustained community spread.

Epidemiologists have started to investigate possible environmental factors that accelerate the spread of SARS-CoV-2 within communities (Sajadi et al., 2020; Bhattacharjee, 2020). A recent paper by van Doremalen et al. (2020) analyzed the aerosol and surface stability of SARS-CoV-2 and compared it with SARS-CoV-1, the most closely related human coronavirus (Wu et al., 2020a). The study found that SARS-CoV-2 can survive up to three days on some surfaces, like plastic and steel, and that aerosol transmission is plausible since the virus can remain viable and infectious in the air for hours. These findings echo those of Chen et al. (2004) on environmental contamination with SARS-CoV-1, and are consistent with evidence for aerosol distribution of SARS-CoV-2 found by Guo et al. (2020), but are inconsistent with the current WHO stance that SARS-CoV-2 is not transported by air. However, the possibility of airborne transmission would call for different mitigation efforts to prevent spreading and is thus an important area of study.

The risk of infection of some airborne viruses has been shown to increase in the presence of ambient fine particles that can stay in the air for long periods, travel far distances, and penetrate deeply into lungs.^1^ One highly contagious airborne disease is caused by the measles virus. Previous studies on disease outbreaks have highlighted that the incidence of measles in China increased 1–3 days after short-term exposure to high concentrations of PM_10_ and SO_2_ Chen et al. (2017b); Peng et al. (2020). In another study, ambient fine particles were found to contribute to the relative risk of influenza transmission in Chinese cities (Chen et al., 2017a) with the most significant effect occurring within a period of 2–3 days.

If air pollution plays a similar role in the incidence of SARS-CoV-2, there should be a positive relationship between confirmed COVID-19 cases and particulate matter concentrations. China ranks among the worst globally in terms of PM_2.5_ concentrations and, within China, the Hubei province is among the more heavily polluted areas (van Donkelaar et al., 2016). The most heavily hit Italian region is the Lombardy area in the northern Po valley, which is among the regions with the worst air quality in Europe. Preliminary findings from Italian researchers started pointing towards a correlation between days of exceeding the limits for PM_10_ and the number of hospital admissions from COVID-19 (Setti et al., 2020; Onufrio, 2020).

Increased air pollution could just reflect the presence of anthropogenic activity which instead explains the patterns. However, that does not explain why COVID-19 cases are not increasing rapidly in every densely populated area.

To investigate this further, the current paper looks at confirmed cases and COVID-19 related hospital admissions in 355 municipalities in the Netherlands and uses regression techniques to investigate correlations between COVID-19 case data and particulate matter concentrations, controlling for a variety of demographic characteristics and data on health related pre-conditions. The analysis finds that PM_2.5_ is a highly significant predictor of both the number of confirmed COVID-19 cases and the number of related hospital admissions per 100,000 inhabitants.

The analysis suggests that the association between air pollution and case incidence is robust to proxies for worse respiratory health and symptom severity. The findings are also robust to other important control variables and different regression specifications that allow for spatial dependence, nonlinearity, alternative error distributions and outlier treatment. Results are obtained with ground-measurements and satellite-derived PM_2.5_. Analyzing COVID-19 data from alternative dates resulted in similar conclusions.

The remainder of this paper is organized as follows. Section 2 visually inspects several available confirmed case maps and discusses the spatial distribution. Section 3 introduces the data used for analysis. Section 4 presents regression results and discusses several of the estimates. Section 5 concludes.

## 2. Spatial Distribution of COVID-19: Country Examples

Suggestive evidence that the spatial distribution of COVID-19 cases is not purely random and might be related to environmental factors can be found by exploring several maps of confirmed cases. A few easily accessible fine resolution maps are presented below, in particular for the Netherlands, Germany, Spain and Italy. The data for the Netherlands is taken from the Dutch National Institute for Public Health and Environment (RIVM).^2^The data for Germany is from the Robert Koch Institute.^3^ The data for Italy can be viewed via a live dashboard, ^4^ and the raw data is well organized and available on a github page.^5^ The Spanish data was taken from this link.^6^

A number of features of the spatial distributions are striking. First, there is a strong spatial correlation visible in all four countries, which is to be expected for a virus that spreads by human contact. It is intriguing, however, that the highest case density in the Netherlands is in Brabant, the southeastern part of the country, while major cities like Amsterdam and Rotterdam are in the west part of the country where the case density is lower. While Brabant is not the most populous province, it accounts for the highest contribution to nation-wide industrial GDP. Within the province, the sub-region Zuidoost-Noord-Brabant produces the highest contribution to industrial GDP.^7^ This area approximately spans the COVID-19 case cluster that can be seen on the map.

**Figure 1:**
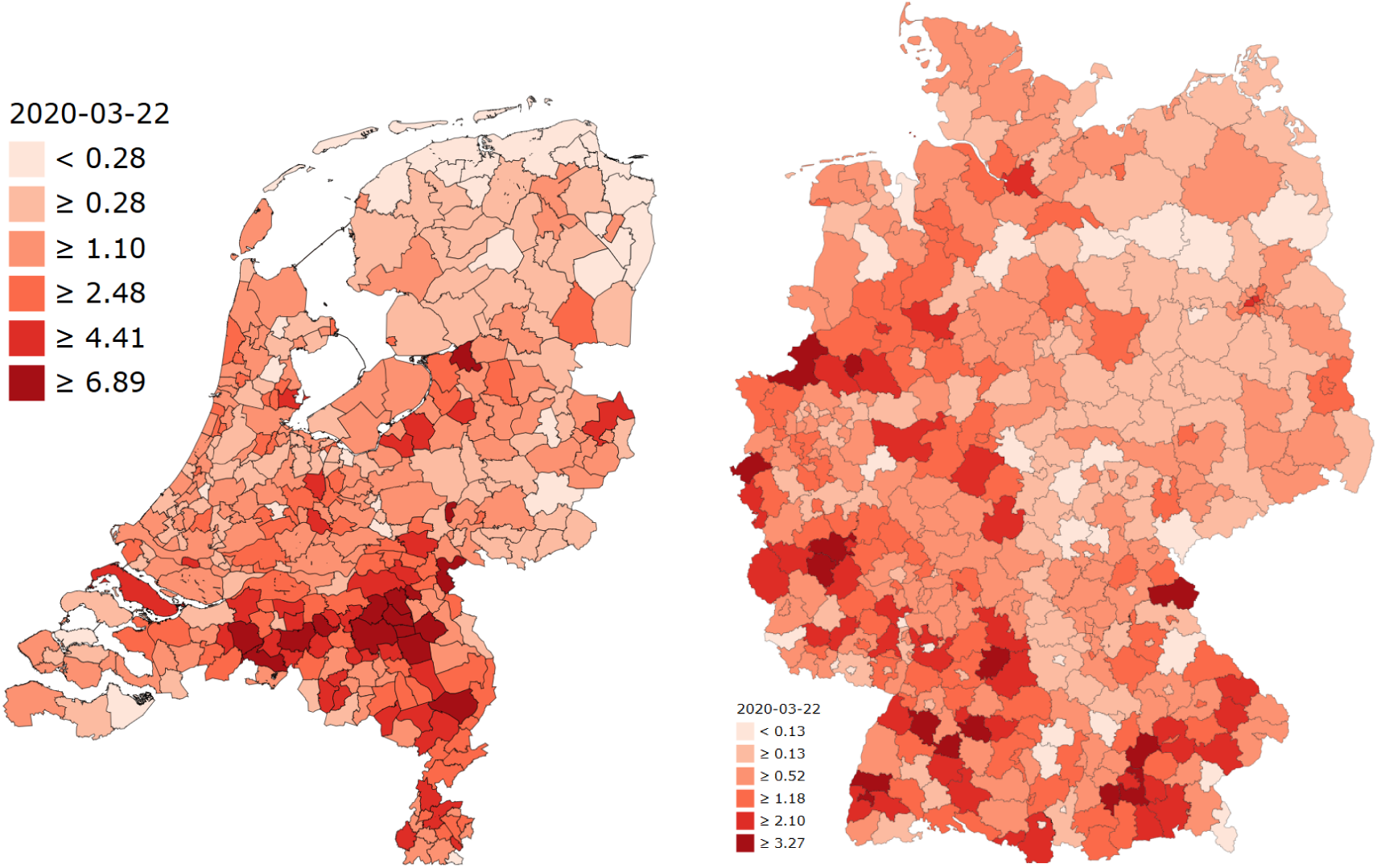
Distribution of COVID-19 in Netherland and Germany. Confirmed cases per 10,000 inhabitants.

**Figure 2:**
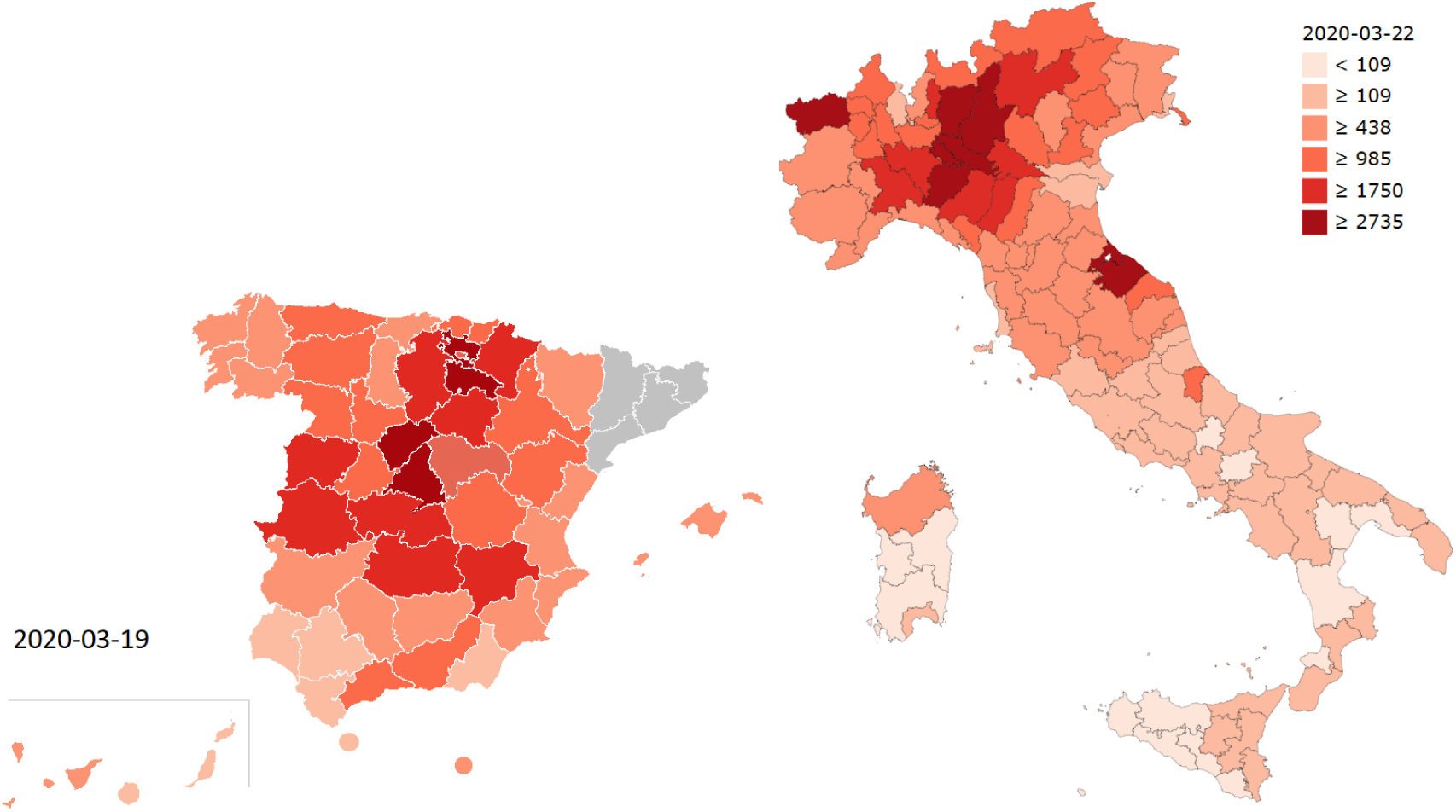
Distribution of COVID-19 in Spain and Italy. Confirmed cases per 1,000,000 inhabitants.

**Figure 3:**
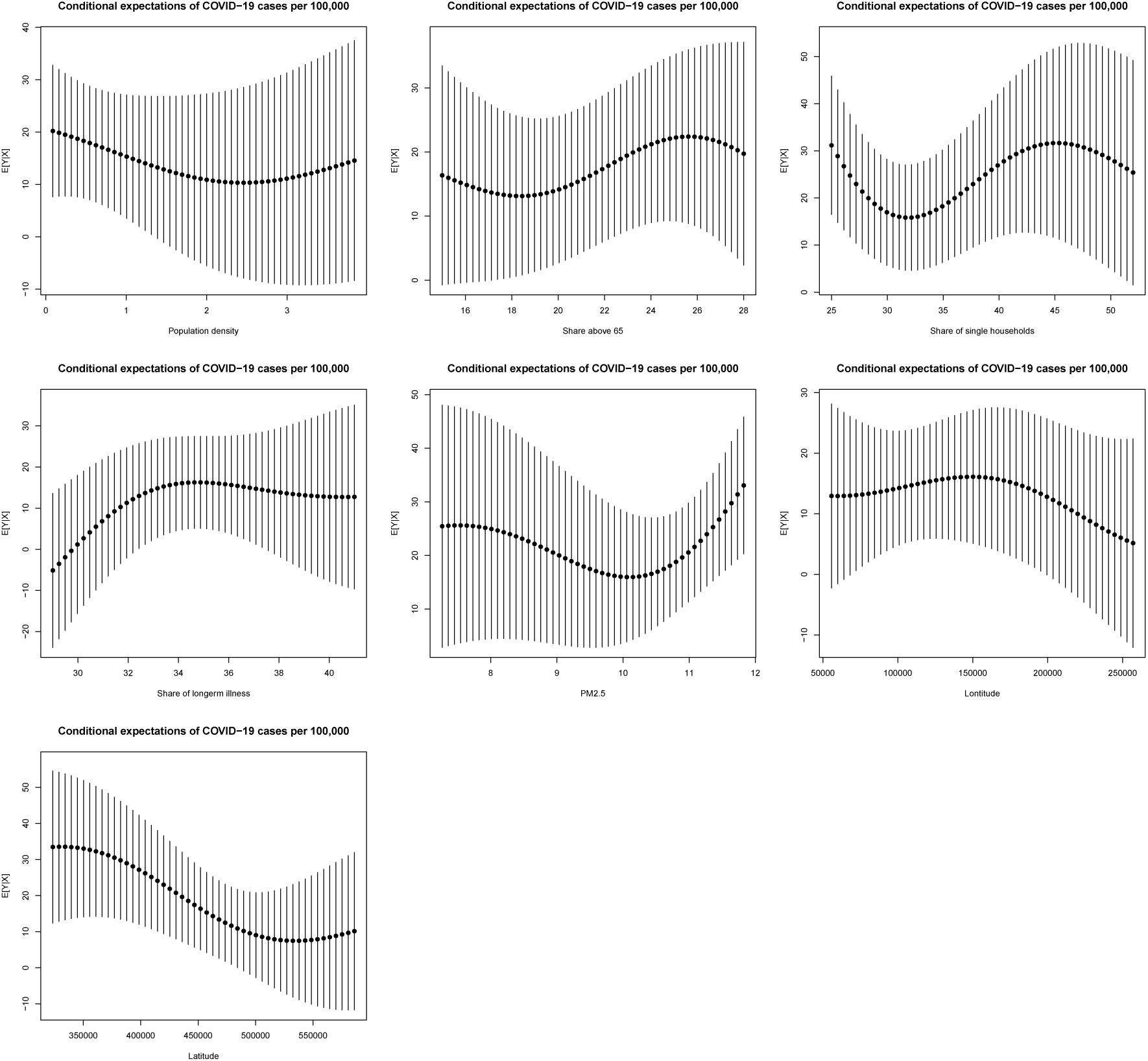
Conditional expectation plots for COVID-19 cases per 100,000 inhabitants.

**Figure 4:**
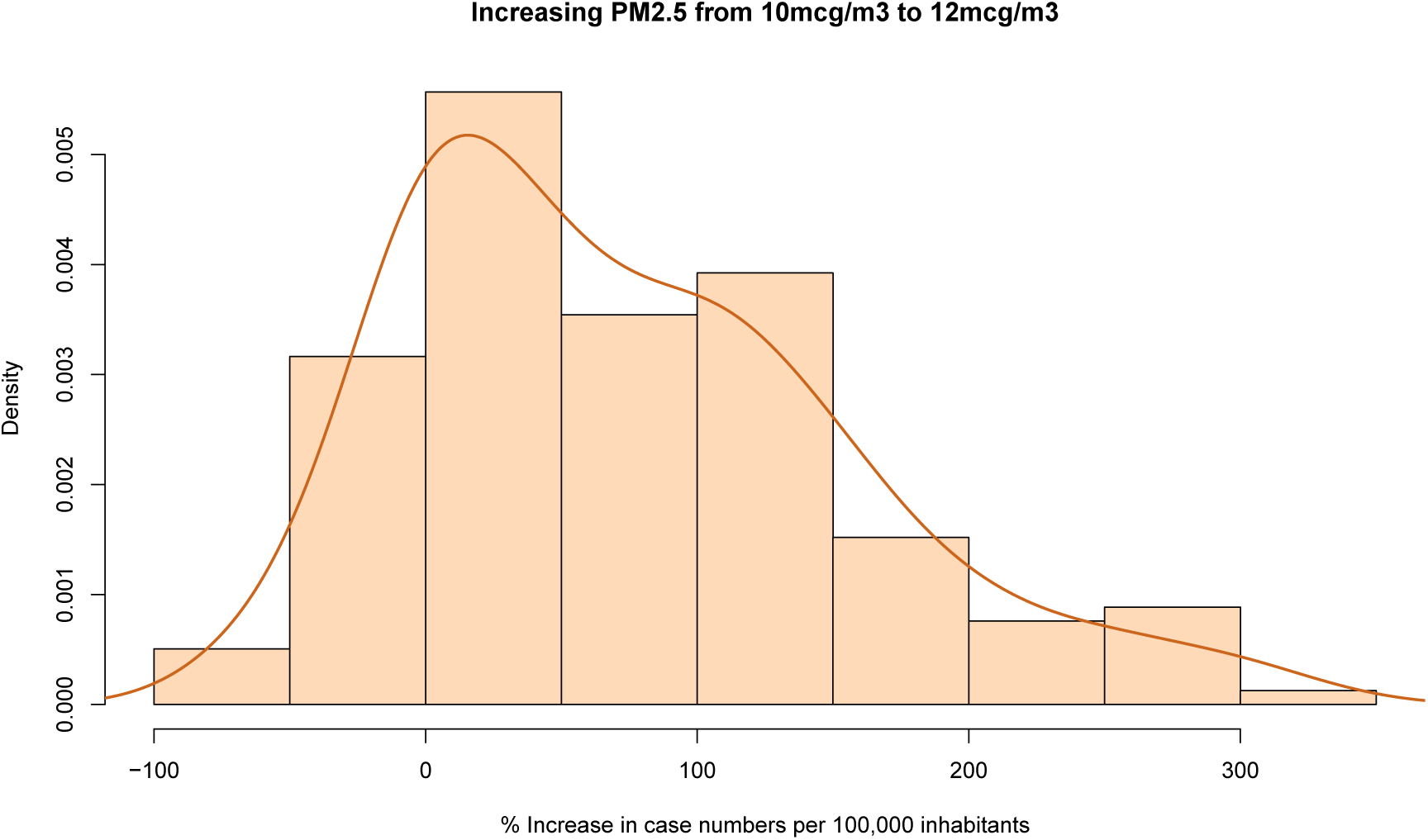
Prediction difference when increasing pollution concentrations (N=158).

In Germany, two areas stand out. First, the western part of the country, near the border with the Netherlands, Belgium, and Luxembourg, has an increased case density. This area (North Rhine-Westphalia, Rheinland-Pfalz and Baden-Württemberg) contains the major industrial regions including the Ruhr area. Second, a cluster of cases can be seen in the south-eastern part of the country near Munich where major automobile industry is found. These areas are also the most populous of the country, which makes it difficult to draw any immediate conclusions about a relationship with air quality.

In Spain, confirmed cases have the highest case density in the capital, Madrid, with an extension into neighboring Sergovia. A second cluster can be also seen northeast of Madrid. Interestingly, Spain’s population density is high along the eastern coast where the case density is lower. This suggests that case incidence in the country does not simply follow population densities, but that other factors play a role.

Finally, in Italy, confirmed cases have the highest case density in the northern part of the country, Lombardy in particular. Without a doubt, Lombardy and the Po valley as a whole has one of the highest concentrations of air pollutants of Europe. Moreover, the case density does not seem to trend strongly with Italy’s population distribution. For example, Italy’s population density is generally high along its coast, and cities like Rome and Naples do not stand out in the map.

Taken together, the maps suggest that that COVID-19 incidence clusters spatially and that environmental factors beyond population density may play a role. The analysis in the remainder of the paper confronts the relatively granular Dutch case data with possible predictors that include population density, air pollution, demographic characteristics and health related controls.

## 3. Data

The COVID-19 data is taken from the RIVM.^8^ The first data snapshot includes all confirmed cases as of March 22 (a total of 4,004 with known residence out of 4,157 confirmed cases). A second snapshot of confirmed cases was taken on March 30 and includes 11,258 cases with known residence out of 11,750 confirmed cases. The confirmed COVID-19 hospital admissions are taken from the same source approximately 1 week after the first data snapshot (31 March, a total of 4,562 with known residence out of 4,712 admissions from a total of 12,595 confirmed cases). While some cases are reported immediately, a share of the cases follows a typical delay of up to 1–2 days after the actual case or hospitalization confirmation. Both the confirmed cases, as well as confirmed hospital admissions, are coded by residence (not by hospital addresses).

On March 31, approximately 37% of confirmed cases were also hospital admissions, highlighting that case detection is likely biased toward more severe cases.^9^ Within one week, the number of hospital admissions exceeded the confirmed cases of the previous week, indicating that the time between confirmation and hospitalization likely spans only a few days. Cases are reported to the RIVM by the Municipal Health Service (GGD).The GGD is organized as collaboration between municipalities to provide base level public health service in accordance with country-level legislation on public health. The 355 municipalities are grouped into 25 GGD areas, each covering a population of approximately 600,000 inhabitants. The GGD borders are visible in figure 5 which visualizes the hospital admissions.

**Figure 5:**
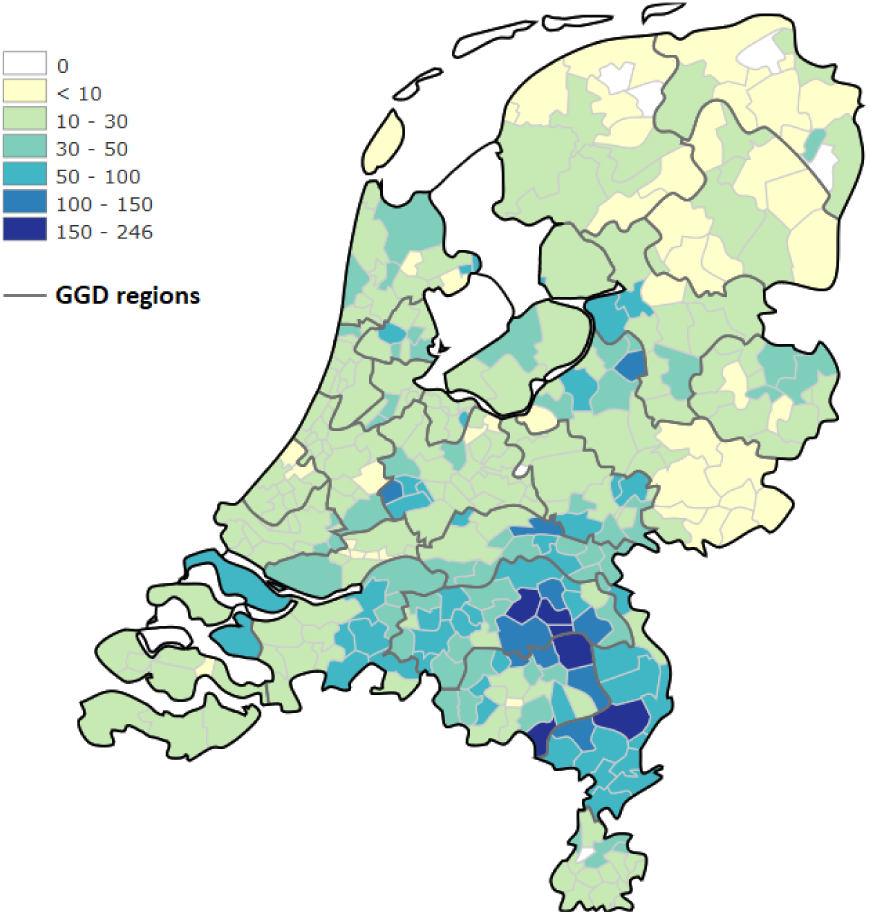
Hospital admissions per 100,000 inhabitants on March 31.

The data is combined with demographic statistics (2019) obtained from the Dutch Central Bureau of Statistics.^10^ The data contains the official population headcount at district level, as well as a number of relevant household characteristics. A number of surveyed health statistics (2016) have been obtained as well from the RIVM (maps can be viewed in the source link).^11^. The data is based on a survey of 457,000 people and includes the share of population in each district with a documented long-term illness (illnesses over 6 months), the prevalence of overweight and obesity, alcohol abuse, smoking and noise due to traffic. Hence the data controls for the presence of possible pre-conditions that make certain populations more vulnerable.

A variety of air pollution data sets exist. For the main analysis, annual average particulate matter concentrations from the RIVM are used to capture long-term exposure (2017, published September 2019).^12^ The data is used by the government for official monitoring in accordance with EU guidelines on air quality monitoring and has a resolution of 25 meter grids. These high-resolution grids are produced by spatial interpolation of ground-measurements. For this analysis, the grids have been averaged to the municipality level. The spatial distribution of pollution has remained relatively stable in recent years. The intensity of air pollution has gradually gone down since 2013, though the difference between the 2017 and 2015 data is relatively small. This suggests that the spatial variation of the 2017 data is still relevant to analyze the role of long-term pollution exposure in the current situation. The temporal lag in the pollution data also ensures that there is no endogeneity due to feedback between case incidence and changes in pollution levels that follow lock-down policies.

To test whether the main findings of the analysis generalize to other pollution data sets, a second analysis presented in the appendix uses the coarser grids from the global PM_2.5_ data set of van Donkelaar et al. (2016). The main conclusions of the analysis do not change when this alternative pollution data set is used, and since this data is mainly satellite-derived, it may be used in other countries where detailed PM_2.5_ measurements are not easily available. Figure 6 visualizes the spatial distribution of the main PM_2.5_ and PM_10_ statistics. Table 3 summarizes the full set of covariates used in the analysis.

**Figure 6:**
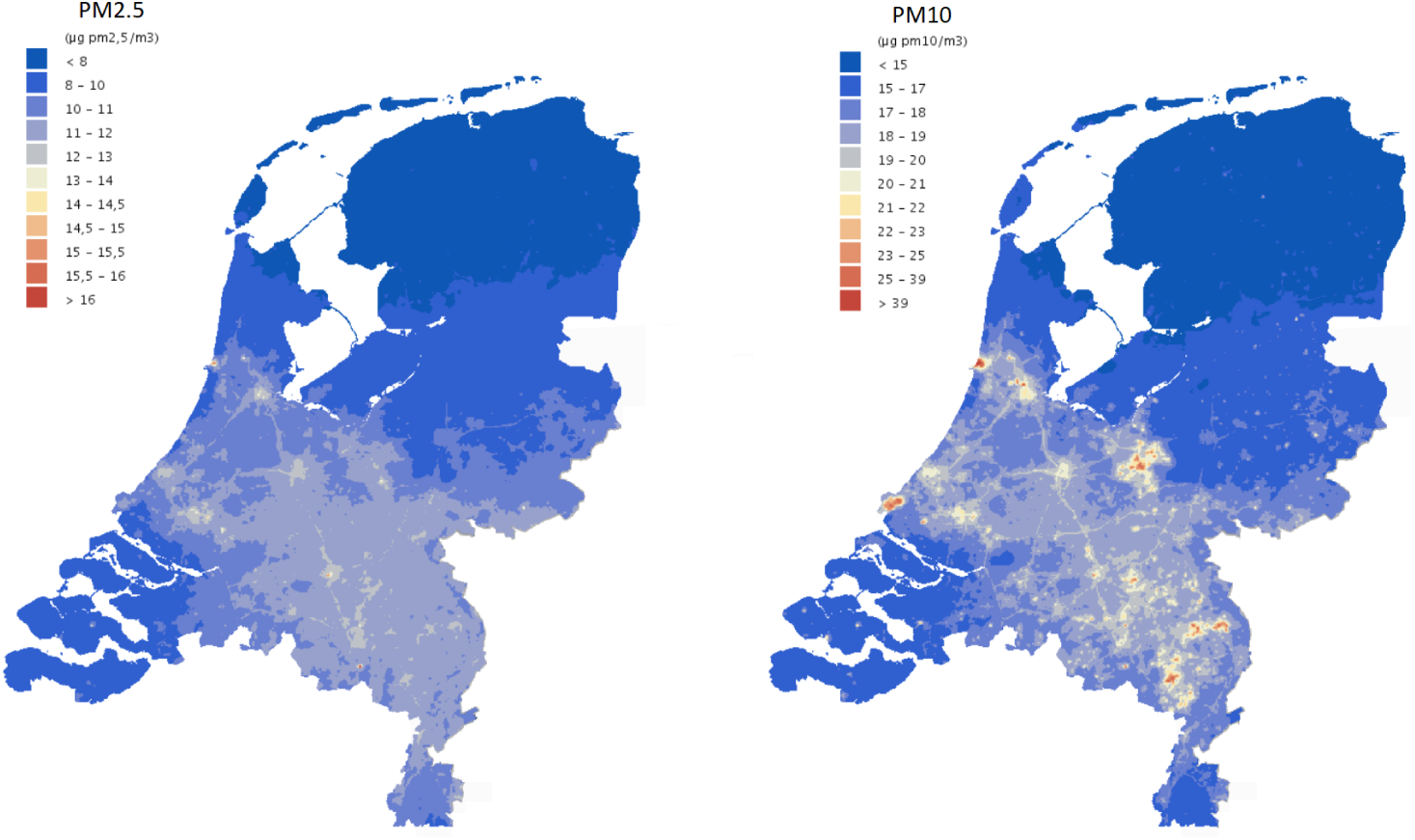
Particulate matter concentrations (2017).

**Table 3:** Data descriptives.

| Variable | mean | sd | median | min | max |
| --- | --- | --- | --- | --- | --- |
| March 22 confirmed cases per 100,000 | 24.79 | 31.62 | 16.00 | 0.00 | 349.50 |
| March 30 confirmed cases per 100,000 | 70.84 | 63.77 | 54.20 | 0.00 | 565.70 |
| March 31 hospital admissions per 100,000 | 29.36 | 30.81 | 20.50 | 0.00 | 237.20 |
| March 22 confirmed case counts | 11.41 | 21.66 | 6.00 | 0.00 | 188.00 |
| March 22 case hospitalization rate | 141.15 | 102.67 | 107.05 | 0.00 | 804.92 |
| March 30 case hospitalization rate | 41.39 | 19.23 | 41.05 | 0.00 | 100.00 |
| Population density (thousands per sqkm) | 0.88 | 1.04 | 0.46 | 0.02 | 6.52 |
| Share of male population | 0.51 | 0.01 | 0.50 | 0.47 | 0.57 |
| Share from 14 to 24 | 11.72 | 1.65 | 12.00 | 9.00 | 23.00 |
| Share from 25 to 44 | 21.96 | 2.84 | 22.00 | 14.00 | 36.00 |
| Share from 45 to 64 | 29.32 | 2.22 | 30.00 | 19.00 | 34.00 |
| Share above 65 | 21.24 | 3.29 | 21.00 | 9.00 | 32.00 |
| Share of unmarried | 44.53 | 4.19 | 44.00 | 37.00 | 65.00 |
| Share of single households | 32.76 | 6.32 | 31.00 | 20.00 | 60.00 |
| Share of households without children | 32.02 | 3.56 | 32.00 | 20.00 | 40.00 |
| Average household size | 2.26 | 0.18 | 2.30 | 1.70 | 3.30 |
| Share of western immigrants | 8.61 | 4.46 | 8.00 | 2.00 | 47.00 |
| Share of non-western immigrants | 7.43 | 5.93 | 5.00 | 1.00 | 39.00 |
| Share of water surface | 0.11 | 0.17 | 0.04 | 0.00 | 0.94 |
| Share with long-term illness | 34.11 | 2.91 | 34.00 | 27.00 | 45.00 |
| Share of overweight | 51.01 | 3.68 | 51.00 | 37.00 | 61.00 |
| Share exposed to noise above 50kmh | 3.03 | 1.38 | 3.00 | 0.00 | 8.00 |
| Share exposed to noise below 50kmh | 4.84 | 2.00 | 5.00 | 1.00 | 13.00 |
| Share with obesity | 14.46 | 2.17 | 14.00 | 9.00 | 22.00 |
| Share of non-heavy drinkers | 39.94 | 4.87 | 40.00 | 30.00 | 58.00 |
| Share of smokers | 19.64 | 2.72 | 19.00 | 14.00 | 31.00 |
| Mean PM<sub>2.5</sub> | 10.22 | 1.33 | 10.69 | 6.95 | 12.04 |
| Mean PM<sub>10</sub> | 17.28 | 1.61 | 17.74 | 13.60 | 21.09 |
| Mean Van Donkelaar PM<sub>2.5</sub> | 14.60 | 1.77 | 15.01 | 10.15 | 18.05 |

## 4. Results

The analysis is organized into two main investigations and a set of robustness analyses. First, section 4.1 analyzes the confirmed cases per 100,000 inhabitants using linear models that account for possible spatial autocorrelation and residual dependence. Section 4.2 analyzes the data nonlinearly, allowing parameter estimates to vary across locations and levels in the data. Additional results are included in the appendix, section 6.2. In particular, the robustness of the results is diagnosed by using alternative measures of incidence, a different source of pollution data, and alternative distributional assumptions.

## 4.1 Linear analysis of March 22 cases per 100,000 inhabitants

To analyze the relationship between spatial variation in particulate matter concentrations and COVID-19 incidence, a number of regressions are estimated that control for possible spatial autocorrelation (Anselin, 1988).^13^ Importantly, the spread of SARS-CoV-2 manifests itself in hot-spots that result from contact with infective subjects from areas that are in close proximity, and it can show strong geographical patterns that are not structurally related to air pollution levels. The fact that the infection started at different times in different areas together with the exponential and geographical nature of case spread, may lead to spurious associations between the spatial distribution of case hot-spots and pollution levels, particularly if the initial cases occurred in polluted regions by mere chance and then spread to nearby regions.^14^

To account for the issue, spatial models include neighboring values of the dependent variable and/or residuals as additional variables. These spatial averages control for the clustering that results from geographical spillovers.^15^ These models can be understood as spatial equivalents to the models that are commonly used to analyze time series in which observed values are in part explained by recent observations. While the household composition and population density terms capture more dense social links, the spatial regression components capture the likelihood of contact with infective subjects. In particular, within a hot-spot, neighboring areas have high numbers of cases per 100,000 inhabitants, and the spatial regression terms capture the increased likelihood of having contact with infective subjects within the region. The important empirical question that these models thus seek to answer is whether pollution and case incidence are associated after controlling for the geographical relationships in disease spread.

First, Model 1 estimates a linear regression using all 22 covariates and possible confounders of interest that are summarized in table 3. These include population density, gender, age groups, marital status and household composition, the share of migrants, as well as several population health indicators. Particularly the health indicators are important because PM_2.5_ is known to affect population health. This may result in important pre-conditions that lead to more severe COVID-19 disease. Pre-conditions captured by the data include the share of population with a long-term illness (including asthma), the share of people that smoke and admit to guidelines on alcohol use, the share of people diagnosed with obesity or overweight, as well as variables on populations exposed to traffic noise.

It is well known that models with a high number of variables can over-fit data sets that contain only a modest number of observations. Model 2 estimates the same linear regression but uses step-wise variable selection following the AIC, Model 3 uses the selected variables in a spatial error model that controls for spatial dependence in the residuals (*λ* parameter), Model 4 estimates a spatial autoregressive model that allows for dependence on neighboring observations (*ρ* parameter), Model 5 allows for unique spatial autocorrelation and spatial residual correlation parameters. For compactness, table 1 only lists Model 1 estimates for variables selected by the AIC, even though all regressors are included. Finally, PM_10_ correlates (.95) strongly with PM_2.5_ and the AIC favored PM_2.5_. Replacing it with PM_10_ in the regressions below led to a small deterioration in measures of fit, indicating that PM_2.5_ is a statistically preferred predictor, although the main conclusions do not depend on this. For simplicity, the focus remains on the PM_2.5_ data.

**Table 1:** Dependent Variable: Confirmed COVID-19 cases per 100,000 inhabitants.

| Variable | Model 1 | Model 2 | Model 3 | Model 4 | Model 5 | Model 5b |
| --- | --- | --- | --- | --- | --- | --- |
| (Intercept) | -359.58<br>(218.20) | -402.51***<br>(80.61) | -202.58***<br>(76.06) | -200.63***<br>(64.11) | -207.50***<br>(74.59) | -185.43***<br>(71.75) |
| Population density | -6.28**<br>(2.58) | -6.54***<br>(2.38) | -0.03<br>(2.05) | -1.10<br>(1.87) | -0.48<br>(2.06) | -0.48<br>(1.94) |
| Share 25 to 44 | 3.55<br>(2.17) | 2.62*<br>(1.34) | -0.80<br>(1.05) | 0.47<br>(1.05) | -0.37<br>(1.07) | -.41<br>(1.02) |
| Share above 65 | 5.58*<br>(2.26) | 4.22***<br>(1.27) | 2.12**<br>(1.03) | 2.14*<br>(1.00) | 2.08**<br>(1.05) | 1.07*<br>(1.00) |
| Share unmarried | 4.94***<br>(1.33) | 4.78***<br>(0.97) | 4.01***<br>(0.96) | 3.28***<br>(0.78) | 3.83***<br>(0.94) | 3.59***<br>(0.89) |
| Share single household | -4.02***<br>(1.47) | -2.17***<br>(0.57) | -1.62***<br>(0.49) | -1.50***<br>(0.45) | -1.59***<br>(0.49) | -1.70***<br>(0.46) |
| Share non-western immigrants | -1.32**<br>(0.57) | -1.23**<br>(0.48) | -0.57<br>(0.43) | -0.78**<br>(0.38) | -0.77*<br>(0.42) | -0.58<br>(0.40) |
| Share of water surface | 17.58<br>(11.18) | 16.11<br>(10.28) | 11.30<br>(9.62) | 13.56*<br>(8.07) | 13.53<br>(9.21) | 11.53*<br>(8.71) |
| Share with long-term illness | 1.10<br>(1.00) | 1.19<br>(0.76) | 0.41<br>(0.92) | 0.72<br>(0.60) | 0.64<br>(0.79) | 0.97<br>(0.74) |
| Case severity |  |  |  |  |  | -0.065***<br>(.01) |
| Mean $PM_{2.5}$ | 10.17**<br>(4.66) | 10.84***<br>(1.48) | 6.21***<br>(2.82) | 3.52***<br>(1.22) | 4.91**<br>(2.44) | 4.47***<br>(2.31) |
| $\lambda$ | | | 0.71***<br>(0.26) | | 0.42*<br>(0.26) | 0.39<br>(0.25) |
| $\rho$ | | | | 0.68***<br>(0.25) | 0.43<br>(0.25) | 0.50**<br>(0.21) |
| R2 | 0.24 | 0.22 | 0.52 | 0.51 | 0.50 | 0.55 |
| AICc | 3414.85 | 3391.40 | 3274.36 | 3274.12 | 3269.31 | 3237.91 |

Standard Errors in parenthesis, significance levels as: \*\*\* $p < 0.01$ , \*\* $p < 0.05$ , \* $p < 0.1$

In the non-spatial regressions, the correlation with population density is negative and significant, suggesting that the case density is on average lower in densely populated areas. This could reflect mis-specified scaling. However, in the models that control for spatial clustering, population density is not significant. This suggests that, after controlling for spatial clustering, the spatial variation in case density is not related to population density patterns. Instead, the share of unmarried and the share of single households, which relate to the number of households in a given population and the type of social networks that they have, are significant regressors. The estimates suggest that instead of the total number of people a person can have contact with within an area (population density), infection risk is determined by the number of people a person is likely to interact with (determined by marital status and household type), together with the average share of infected inhabitants in the wider region (spatial components). This is a plausible result. To simplify the multi-dimensional relationship between case densities and social interaction, regressing single household shares on unmarried population shares shows that on average, a 1% increase in the first is associated with a 0.74 % increase in the latter, suggesting that on average the case densities increase when there are more households in an area.

Importantly, across all regression specifications, the coefficient for PM_2.5_ is positive and highly significant in the presence of controls, and also in the specifications that control for spatial residual trends and the rate of infective subjects in nearby areas. Combined, the regressions thus provide strong evidence that PM_2.5_ plays a role in COVID-19 case incidence that cannot be attributed to demographics or health pre-conditions. In particular, the estimate of 10.84 in Model (2) suggests that, on average, cases per 100,000 inhabitants grow by approximately 21.68 ~ 22 when concentrations increase from 10mcg/m3 to 12mcg/m3. This corresponds to slightly less than a 100% increase given that the average municipal case density in the data is 24.79 ~ 25 per 100,000 inhabitants. Note that the direct elasticity is lower in the spatial models (3–5), but the net impacts need to be multiplied by spatial spillover effects. Spatial spillovers *ρ* and spatial correlation in the residuals *λ* are both significant and have a positive sign highlighting that spatial spillovers further add to local effects. For example, evaluating the prediction difference of the spatial autoregressive model (4) at PM_2.5_ levels of 10mcg/m3 and 12mcg/m3, suggests a very similar increase in case incidence of 22.08 ~ 22 per equal number of inhabitants.

Across the regression specifications, it is found that the health indicators have no significant linear relationship with confirmed case incidence. Only the share of population with a long-term illness was kept in the model with the lowest AIC, but its effect is not significant in any of the regression specifications. Going from Model 1 to Model 2, it can be seen that the parameter estimate for PM_2.5_ varies little after dropping the majority of health data controls, suggesting that the association between case incidence and pollution is not heavily impacted by adding or removing available data on possible pre-conditions. It is however important to ensure that the association between case incidence and pollution concentrations is not in fact driven by worse respiratory health in polluted areas. If worse respiratory health and aggravated symptoms in polluted areas are the main channels of action, higher COVID-19 case hospitalization rates should also be expected in these locations. For this reason, the percentage of the confirmed cases that resulted in hospital admission one week later (March 22 cases / March 31 hospital admissions times 100) was calculated as proxy for case severity. In 29 areas with no confirmed cases where hospitalizations occurred within a week, a value of 100 is assigned. In 9 areas where none of the confirmed cases resulted in hospitalization, a value of 0 is assigned.^16^ Model 5b adds this additional proxy variable and finds that it is highly significant. The overall model fit improves as indicated by the AICc and R2. The estimate for PM_2.5_ remains relatively unchanged and significant. While one would expect that case severity contributes to higher cases, as increased symptom severity may lead to higher case detection, the result suggests otherwise. One explanation is that high hospital admission occurred in areas with a weak case detection policy. In particular, if the disease goes unnoticed for long, the number of terminally ill patients can grow because they do not receive appropriate treatment in time. In this case, low case numbers can coincide with high hospital admission numbers. To investigate further whether the proxy captures a valuable signal related to symptom severity, appendix section 6.2.5 provides additional results that try to explain the case severity proxy using the other available predictors. These additional results find that age, male gender, and the share of population with overweight are positively associated with increased case hospitalization rates. This is in line with earlier identified risk groups (Ruan, 2020), suggesting that the proxy does capture a relevant case severity signal.

Taken together, the evidence suggests a significant positive relationship between case density and PM_2.5_ concentrations. However, there are still some limitations to the basic regression results presented here. The standard linear regression model may not be perfectly suitable for modeling the number of cases per 100,000 inhabitants due to the non-negative nature of the data and a right skew in the case density distribution. Strong violations of the correct-specification assumption can result in biased estimates, for instance because the models assess linearity on an additive scale while the phenomenon is multiplicative. Instead of assessing the data on the original scale as a multiplicative error model with a changing variance function, this issue is often addressed by rewriting the model as an additive error model on the log-scale with constant variance. This is appropriate as long as the log transformation is appropriate to normalize the data. To assess whether the simple estimations presented here are prone to a strong bias, section 6.2.2 investigates the residual distribution and re-analyzes the data using a log-type power transformation from a family of functions that allows for zeros. The results highlight that when the data is appropriately scaled and multiple diagnostics confirm that the Normality assumption is in fact valid, PM_2.5_ is still a highly significant positive predictor of case densities. Earlier studies on the role of ambient fine particles in the transmission risk of airborne disease have instead relied on Poisson-type regressions using count data. While these regressions are not entirely appropriate as they do not account for the highly significant geographical relationships in the data, section 6.2.1 presents Poisson-type results that allow for over-dispersion to show that the main conclusions are also robust to this specification choice.

### 4.2 Nonlinear analysis of March 22 cases per 100,000 inhabitants

Instead of working on a transformed scale to address some of the highlighted issues, it is also possible to tackle the problem nonlinearly on the original scale. This might also lead to interesting results on important thresholds in the data. In particular, one might expect particulate matter to only contribute to COVID-19 incidence after concentrations surpass a certain critical threshold, or expect pollution dependencies to vary with unobserved weather variables including humidity and temperature (Chen et al., 2017b,a; Peng et al., 2020). In a recent study, Sajadi et al. (2020) have already shown that there could be a relationship between COVID-19 incidence and climatic conditions. Some COVID-19 related climatic zones are mapped by the Copernicus Earth Observation Programme, see (Copernicus Climate Change Service, 2020), and these put all but a select few municipalities analyzed in this study in the same zone. For this reason, one should expect that if the relationship between pollution and COVID-19 incidence varies regionally, it does so with a reasonable smoothness.

Additional results below are obtained using non-parametric penalized kernel regression following (Hainmueller and Hazlett, 2014; Andrée et al., 2019). The estimates provide observation-level marginal coefficients that allow for nonlinearity conditional on levels in the data. Longitude and latitude have been added as additional controls, which allows the model to capture spatial trends in line with a spatial residual component. However, this time it also allows the model’s parameters to vary across spatial gradients in unobserved components, such as related to weather. The model nests a linear model, specifically, higher levels of regularization result in linearized relationships. Evidence that the relationship with air pollution is nonlinear is strengthened by using Model (2) and re-estimating it after applying a third-order Taylor approximation to the PM_2.5_ measurements. Calculating an auxiliary test statistic for the significance of the second and third terms overwhelmingly supports nonlinearity, a Likelihood Ratio obtains a *p*-value below 0.001 (statistic of 15.18 on 2 degrees of freedom).

The fit of the nonparametric model is tuned using standard cross-validation procedures and out-of-sample prediction performance was estimated using 10-fold, repeated twice, cross-validation. To keep the flexibility of the models at a manageable level given the small number of observations, only a few predictors are used. In particular, the significant predictors from the final model (5) are taken, the share of unmarried is dropped as the model can now estimate nonlinear dependence on the share of single households, the share of long-term illness and population density are added back in because they remain of particular interest. The share of non-Western immigrants was dropped because it was insignificant and dropping it did not negatively impact the cross-validation results. Finally, the share of population in the 25 to 44 years group was dropped because estimating nonlinearly on only the share of population above 65 resulted in better fit.

Table 2 presents the estimation results. In it, Avg. takes the average across all the marginal coefficients and q.25-q.75 give the quantiles as an indication of parameter heterogeneity. To understand the shape of the nonlinearity, figure 3 plots conditional expectations across the range of values in the covariates. The values are produced by fixing all covariates, except the one of interest, at mean values, and plotting the model predictions and their standard errors across the .025th percentile to the .975th percentile values of inputs.

**Table 2:**
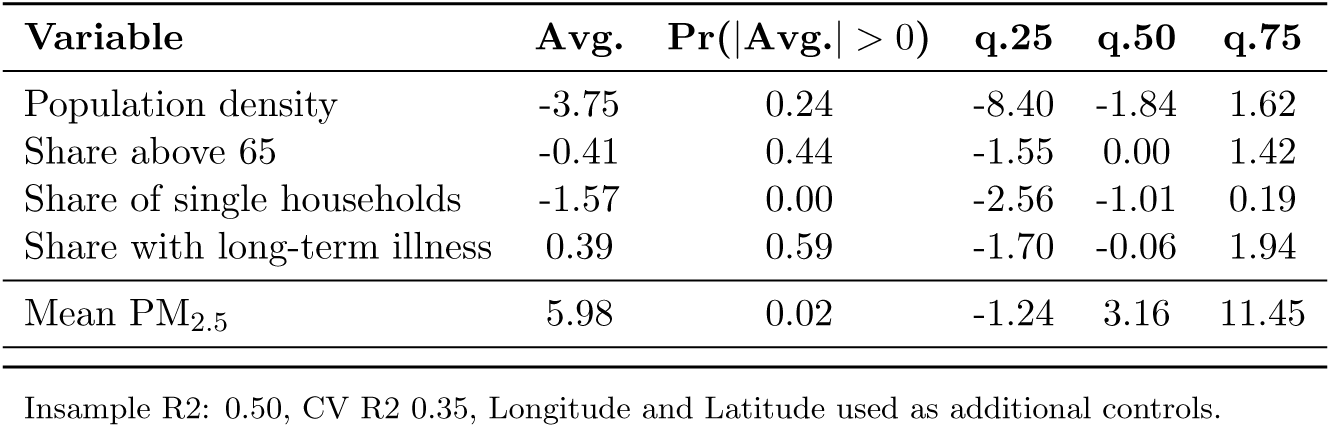
Dependent Variable: Confirmed COVID-19 cases per 100,000 inhabitants.

| Variable | Avg. | Pr(\|Avg.\| > 0) | q.25 | q.50 | q.75 |
| --- | --- | --- | --- | --- | --- |
| Population density | -3.75 | 0.24 | -8.40 | -1.84 | 1.62 |
| Share above 65 | -0.41 | 0.44 | -1.55 | 0.00 | 1.42 |
| Share of single households | -1.57 | 0.00 | -2.56 | -1.01 | 0.19 |
| Share with long-term illness | 0.39 | 0.59 | -1.70 | -0.06 | 1.94 |
| Mean $PM_{2.5}$ | 5.98 | 0.02 | -1.24 | 3.16 | 11.45 |

Insample R2: 0.50, CV R2 0.35, Longitude and Latitude used as additional controls.

The averages of parameter estimates resemble the results from those obtained with linear regression methods. In particular, the average slope of population density is again negative but not significant, while the increased share of single households provides a stronger signal for increased case densities, particularly in the inner range of values that have denser data coverage (see figure 3). The age group control shows that elderly are more at risk. The estimated relationship with the share of population that has a long-term illness highlights an important threshold. Fewer COVID-19 cases are expected only in areas with very low values for this indicator.

Importantly, after addressing nonlinearity and spatial heterogeneity in parameter estimates, the average slope of PM_2.5_ remains positive and highly significant. The ranges in the quantiles highlight that there is substantial parameter heterogeneity. The nonlinear estimates suggest that at low levels of PM_2.5_, changes in particulate matter concentrations are not associated with significant changes in case incidence. However, after the mean annual concentrations cross the WHO guidelines of 10mcg/m3, the standard errors tighten and the number of expected cases increases sharply. At 12mcg/m3, the expected cases per 100,000 inhabitants are approximately double the numbers expected at 10mcg/m3.

More indication of impacts is approximated by calculating the prediction difference when PM_2.5_ moves from 10mcg/m3 to 12mcg/m3, leaving all other covariates at observed values. This is performed for all areas that have at least already 9mcg/m3 and case numbers within the 25% to 75% quantile range (between 8.3 and 31.7 cases per 100,000). The prediction difference is standardized based on the current actual case numbers and multiplied by 100, thus expressed as a percentage increase with respect to current case numbers. The results in figure 4 highlight the effect heterogeneity, suggesting that the modeled pollution association varies strongly depending on other covariates. Numerical integration under the kernel density suggests 80% of events result in positive increases in case incidence, and of these events the estimated average increase in cases per 100,000 inhabitants is 95% when particulate matter concentrations increase from 10mcg/m3 to 12mcg/m3.

## 5. Conclusion

Research on viral respiratory infections, measles and influenza outbreaks has found that infection risks increase following exposure to high concentrations of particulate matter. This paper investigated the relationship between COVID-19 incidence and exposure to particulate matter in 355 municipalities in the Netherlands. Regression analysis was performed using confirmed cases per 100,000 inhabitants, confirmed COVID-19 related hospital admissions per 100,000 inhabitants, and confirmed case counts as dependent variables.

The study finds that PM_2.5_ is a highly significant predictor of all three indicators of COVID-19 incidence. The findings are robust to outlier treatment and power transforms to normalize data, and are stable across alternative regression specifications that allow for spatial dependence or nonlinearity, and remain significant in the presence of demographic and health controls. Estimates suggest that when annual concentrations cross above the WHO guidelines of 10mcg/m3, the number of expected cases per 100,000 inhabitants doubles as annual concentrations reach 12mcg/m3 all else constant.

While the analysis found that these results are robust to various methodological considerations, it is important to note that testing for SARS-CoV-2 is performed using convenience sampling, which may well vary by area and in time. This may induce biases in the results if the sampling rate is indirectly correlated with pollution levels. However, it is difficult to perceive why sampling should structurally be related to pollution concentrations. Moreover, in light of the rich body of literature on the association between pollution exposure and respiratory tract infection risk, and the plausible parameter estimates with respect to many of the other variables, convenience sampling does not seem to be a more plausible explanation for the results than the findings of the study itself. Moreover, another new study by Wu et al. (2020b) has found evidence for a higher number of confirmed fatal COVID-19 cases per 100,000 inhabitants in the United States, which seems to corroborate the findings on increased hospital admissions and cases per 100,000 inhabitants.

The findings call for further investigation. In particular, the air pollution link should be investigated in multiple countries and for wider ranges of PM_2.5_ concentrations. If the relationship extrapolates to higher concentrations, the implications for developing countries may be severe. In particular, developing countries are highly polluted compared to the levels observed in this study (Andrée et al., 2019) and are already identified as risk areas for COVID-19 spread (Gilbert et al., 2020; Nkengasong and Mankoula, 2020; Martinez-Alvarez et al., 2020). Even though this study was not able to find strong evidence for an impact of PM_2.5_ on case severity, at the high levels of PM_2.5_ in developing countries, more severe impacts on respiratory health may interact with case fatality of SARS-CoV-2. The possible association between pollution and symptom severity will thus be important to revisit, particularly because regional variation in case fatality of closely related SARS-CoV-1 has been associated with air pollution exposure (Cui et al., 2003).

Finally, as more data on COVID-19 spread becomes available, stronger results on the specific effects of short-term air pollution exposure may be estimated. If fine particulate matter plays a significant role in SARS-CoV-2 infection risk, it has strong implications for the mitigation strategies required to prevent spreading.

## 6. Appendix

### 6.1 Data descriptives

### 6.2 Additional analysis results

This section presents a number of additional estimations. In particular, section 6.2.1 presents Poisson-type regressions that use case counts as dependent variables in line with earlier studies on the relationship between air pollution and viral spread, the influence of possible outlier observations or the discussed distributional issues is investigated in section 6.2.2, in section 6.2.3 the main analysis is re-estimated using confirmed cases from March 30 to show that the conclusions are not dependent on the date of the case snapshot, the main analysis is also repeated using confirmed hospital admissions from March 31 in section 6.2.4 to provide further evidence that the conclusions are not dependent on measurement error in the confirmed cases. Section 6.2.5 investigates correlations between covariates and the case hospitalization rates used to proxy for symptom severity. Finally, additional analysis in section 6.2.6 re-estimates the main analysis using alternative satellite-derived PM2_2.5_.

#### 6.2.1 Poisson-type regressions for case incidence

In table 4, the case counts are used in regression instead of case density. Model 8 estimates a standard Poisson regression. Model 9 estimates a Poisson regression with stepwise selection following the AIC. Model 10 presents the same model, allowing for over-dispersion. Finally, Model 11 allows for a zero-inflated Negative Binomial distribution and model 12 performs step-wise AIC under the Negative Binomial.

**Table 4:** Dependent Variable: Confirmed COVID-19 cases.

| Variable | Model 8 | Model 9 | Model 10 | Model 11 | Model 12 |
| --- | --- | --- | --- | --- | --- |
| (Intercept) | -13.75***<br>(2.23) | -13.75***<br>(1.64) | -13.75***<br>(5.12) | -9.89**<br>(4.86) | -4.14<br>(2.72) |
| Population density | -0.35***<br>(0.02) | -0.35***<br>(0.02) | -0.35***<br>(0.08) | -0.30***<br>(0.08) | -0.29***<br>(0.07) |
| Share of male population | -22.38***<br>(1.90) | -23.12***<br>(1.85) | -23.12***<br>(5.77) | -16.07***<br>(4.96) | -13.29***<br>(4.41) |
| Share 14 to 24 | 0.07***<br>(0.02) | 0.07***<br>(0.01) | 0.07<br>(0.04) | 0.04<br>(0.05) |  |
| Share 25 to 44 | 0.16***<br>(0.02) | 0.16***<br>(0.02) | 0.16***<br>(0.05) | 0.17***<br>(0.05) | 0.15***<br>(0.04) |
| Share above 65 | 0.13***<br>(0.02) | 0.13***<br>(0.02) | 0.13**<br>(0.05) | 0.09*<br>(0.05) | 0.05*<br>(0.03) |
| Share of unmarried | 0.14***<br>(0.01) | 0.14***<br>(0.01) | 0.14***<br>(0.04) | 0.11***<br>(0.04) | 0.10***<br>(0.02) |
| Share of household without children | 0.06***<br>(0.02) | 0.06***<br>(0.01) | 0.06<br>(0.04) | 0.04<br>(0.04) |  |
| Average household size | 2.75***<br>(0.44) | 2.58***<br>(0.25) | 2.58***<br>(0.77) | 1.04<br>(0.68) |  |
| Share of western immigrants | 0.03***<br>(0.01) | 0.03***<br>(0.01) | 0.03<br>(0.02) | 0.00<br>(0.02) |  |
| Share of non-western immigrants | 0.03***<br>(0.00) | 0.03***<br>(0.00) | 0.03**<br>(0.01) | 0.02<br>(0.02) |  |
| Share enduring noise above 50km | 0.03*<br>(0.02) | 0.04***<br>(0.01) | 0.04<br>(0.04) | 0.01<br>(0.04) |  |
| Share of obese | 0.06**<br>(0.02) | 0.05***<br>(0.02) | 0.05<br>(0.05) | -0.03<br>(0.04) |  |
| Share of non-drinkers | -0.05***<br>(0.01) | -0.05***<br>(0.01) | -0.05**<br>(0.02) | 0.00<br>(0.01) |  |
| Share of smokers | 0.11***<br>(0.01) | 0.12***<br>(0.01) | 0.12***<br>(0.04) | 0.06<br>(0.04) |  |
| Mean PM<sub>2.5</sub> | 0.52***<br>(0.05) | 0.47***<br>(0.02) | 0.47***<br>(0.06) | 0.45***<br>(0.05) | 0.44***<br>(0.04) |
| R2 | 0.59 | 0.59 | 0.59 | 0.47 | 0.47 |
| AICc | 3995.20 | 3983.77 | 3983.77 | 2242.19 | 2227.05 |

Standard Errors in parenthesis, significance levels as: \*\*\* $p < 0.01$ , \*\* $p < 0.05$ , \* $p < 0.1$

The residuals are checked for spatial autocorrelation, and significant residual clustering was still found. Spatial autocorrelation is not easily addressed in count data with standard regression implementations, hence the results are simply from a mis-specified model. For this reason, the models that relax distributional assumptions (10–12) should provide improved indications of significance, with Model 10 being proffered. The main results of the paper presented in section 4 take spatial processes explicitly into account and are in turn preferred over Model 12.

A few results using the count data echo the main findings. In particular, the slope of population density is negative, indicating that the more populous areas in the Netherlands are more likely to have lower case numbers on average rather than higher. Several of the health indicators are significant, but only in the standard Poisson regression. Allowing for over-dispersion or estimating under the Negative Binomial distribution finds no significant relationship suggesting that these relationships are less robust. In all specifications, the impact of PM_2.5_ remains highly stable and significant. This provides further evidence that confirmed COVID-19 cases are higher in polluted areas and that these conclusions do not dependent on using count or Normal estimation techniques.

#### 6.2.2 Distributional mis-specification and outlier analysis

Since only a modest amount of observations has been used in the analysis, it is important to diagnose whether the estimation result could be heavily impacted by outlier observations. One way to diagnose this is to inspect a Q-Q plot, which compares the standardized residuals to theoretical quantities from the Normal distribution.

Figure 7 highlights that the Normality assumption is not entirely satisfied. Both Model 2 and Model 5 residuals contain outliers, particularly in the right tail of the distribution. In both models, the residuals follow a very similar pattern and the three major outliers that are prevalent on both specifications are Boekel, Uden and Bernheze which are all in the COVID-19 cluster in the province of Brabant. Outliers can be influential in a regression, though they do not necessarily have to be, while other points that lie within a normal range of the model can be influential without being an outlier per se. The impact of outlier observations depends also on the data density in the region around the data point. The Q-Q plots do not inform about whether the identified outlier observations are actually influential in the regression. Figure 8 calculates Cook’s distance, a multivariate measure of influence, and identifies influential data points by evaluating the impact of individual observations on the regression results with respect to the covariates of interest through a leave-one-out procedure.

**Figure 7:**
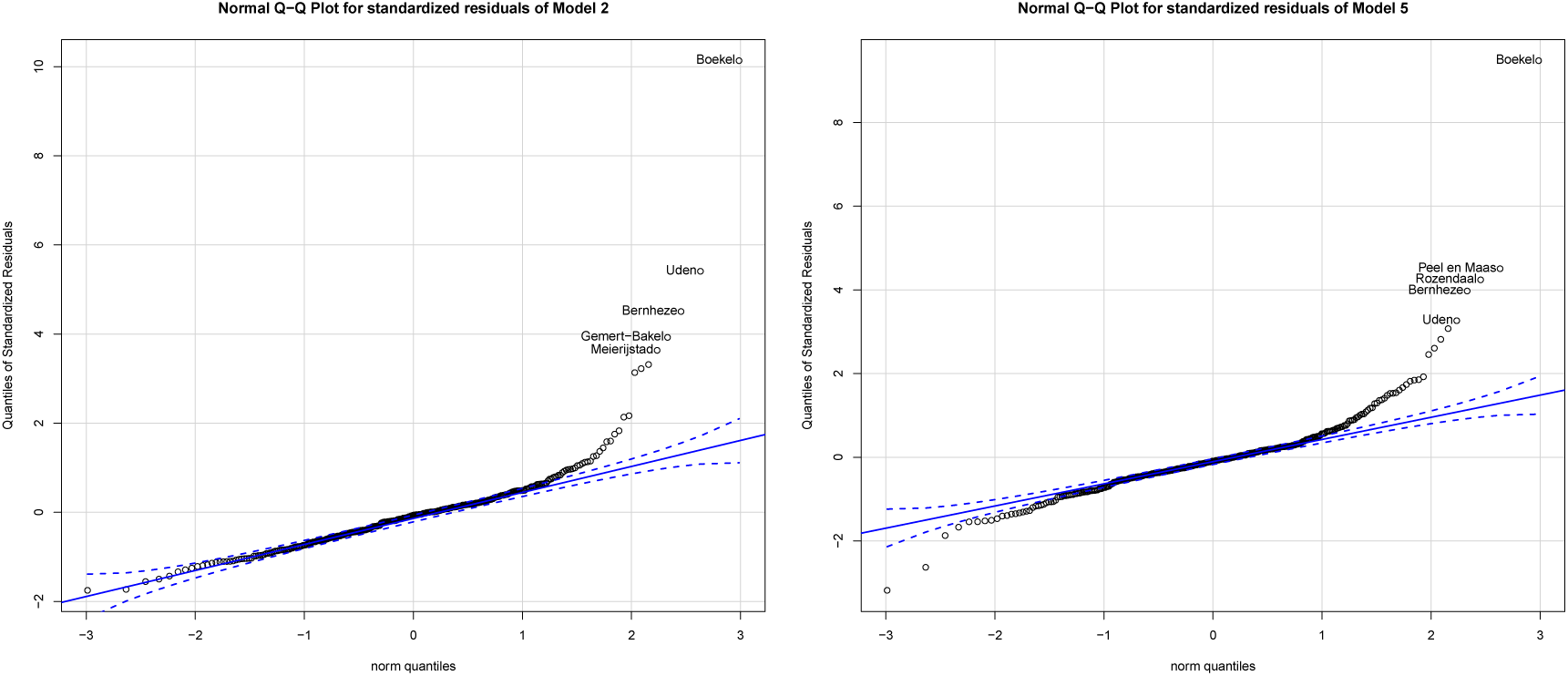
Comparison of residuals to theoretical quantities.

**Figure 8:**
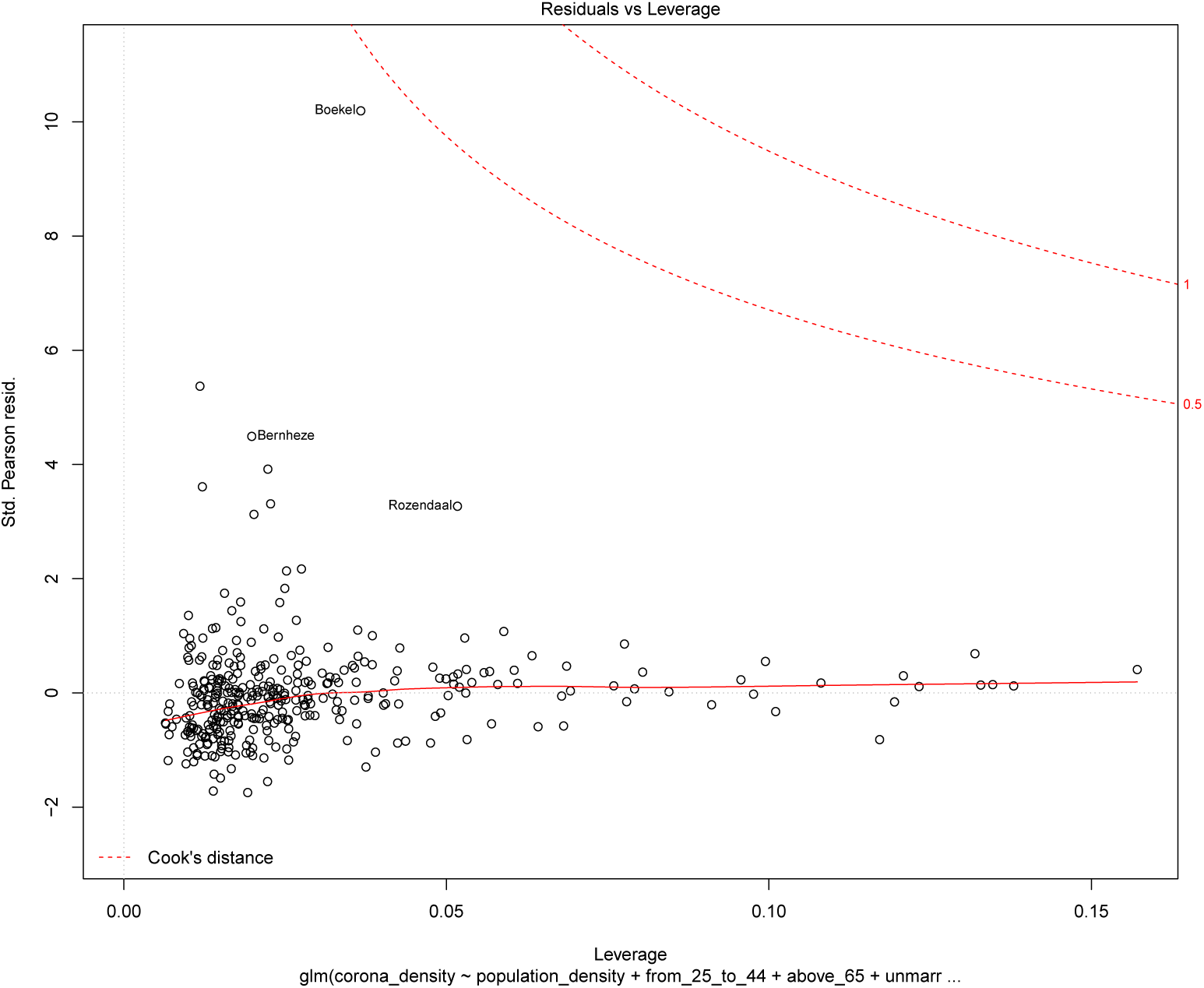
Residuals versus Leverage plot using Cook’s distance.

Figure 8 highlights that Boekel and Bernheze are relatively influential, but not critically. This is reassuring, nevertheless it is important due to the small sample nature of the applications to evaluate whether the identified mild violations have a drastic impact on the estimation results. Two regressions are performed to analyze this. First, Model 5c replaces the dependent values of the 8 observations that have visibly the largest residuals in the Q-Q plot with predicted values from Model 5 and re-estimates the specification. This allows comparing directly how the parameter estimates change when these outlier observations are replaced with values that lie closer to the normal range of the data. It is important to note that if these observations are not outliers in an additive sense, but simply reflect the nature of the data-generating process, then these new estimates have in fact an increased bias resulting from further mis-specification. To evaluate whether outliers can be addressed through model-specification, Model 5d first normalizes the data using a power transformation (Johnson) by finding the transformation that minimizes the *p*-value of a Shapiro test for Normality, and then re-estimates Model 5’s specification on the more Gaussian data. These parameter values cannot be directly compared to the parameter values of Model 5 because the new relationship is nonlinear (logarithmic-type). Nevertheless, Model 5d informs whether the significance of relationships remains intact when the Normality violations are neutralized.

**Table 5:** Dependent Variable: Confirmed COVID-19 cases per 100,000 inhabitants.

| Variable | Model 5c | Model 5d |
| --- | --- | --- |
| (Intercept) | -91.40**<br>(43.93) | -10.75***<br>(2.41) |
| Population density | -0.83<br>(1.27) | -0.16**<br>(0.07) |
| Share from 25 to 44 | -0.16<br>(0.73) | 0.05<br>(0.04) |
| Share above 65 | 0.67<br>(0.68) | 0.10**<br>(0.04) |
| Share of unmarried | 1.61***<br>(0.52) | 0.10***<br>(0.03) |
| Share of single households | -0.56*<br>(0.30) | -0.05***<br>(0.02) |
| Share of non-western immigrants | -0.55**<br>(0.25) | -0.02<br>(0.01) |
| Share of water surface | 9.33*<br>(5.31) | 0.68**<br>(0.31) |
| Share with long-term illness | 0.29<br>(0.37) | 0.01<br>(0.02) |
| Mean PM<sub>2.5</sub> | 2.67***<br>(0.89) | 0.45***<br>(0.04) |
| $\lambda$ | -0.22<br>(0.16) | -0.05<br>(0.19) |
| $\rho$ | 0.73<br>(0.07) | -0.06<br>(0.17) |
| R<sup>2</sup> | 0.54 | 0.31 |

Standard Errors in parenthesis, significance levels as: \*\*\* $p < 0.01$ , \*\* $p < 0.05$ , \* $p < 0.1$

Inspecting the new Q-Q plots in figure 9 highlights that Model 5c is still prone to distributional mis-specification. This also suggests that the outliers whose values are now replaced with values closer to the normal range of the data are not necessarily outliers in an additive sense, but simply reflect the exponential nature of the data. From that regard, Model 5d is preferred, as it applies a suitable exponential transformation that clearly neutralizes any outlier or non-Gaussian behavior. In both models, the relationship with PM_2.5_ remains significant and positive hence the conclusion is that the main findings of the analysis are not sensitive to outliers.

**Figure 9:**
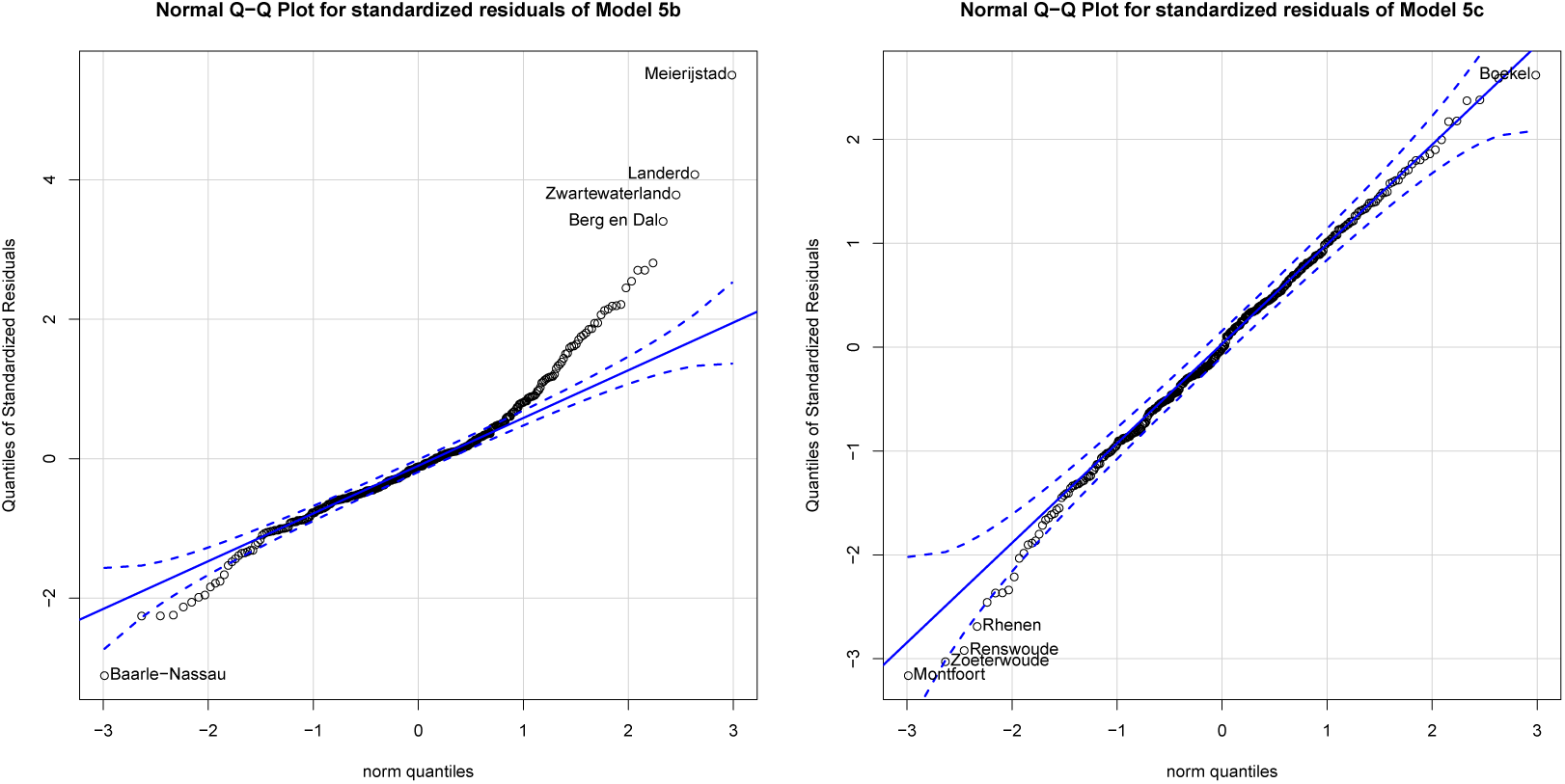
Comparison of residuals to theoretical quantities.

#### 6.2.3 Re-estimation using March 30 confirmed cases per 100,000 inhabitants

Model 5e, in table 6 below, re-estimates the step-wise AIC regression and then uses the covariates in the same specification as Model 5 using confirmed cases from March 30. This is to evaluate whether the relationship with PM2_2.5_ is robust to using data from a different date. The correlation between confirmed cases per 100,000 inhabitants on March 22 and March 30 is approximately .90. It is clear from the estimation results that using the newer data does not alter the main conclusions. In particular, similar covariates are preferred by the AIC and the parameter estimate for PM_2.5_ increased in value and remains significant at the highest level.

**Table 6:** Dependent Variable: Confirmed COVID-19 cases per 100,000 inhabitants using March 30 cases.

| Variable | Model 5e |
| --- | --- |
| (Intercept) | -374.01***<br>119.87 |
| Population density | -2.28<br>(3.54) |
| Share from 25 to 44 | 3.12<br>(1.99) |
| Share above 65 | 4.79***<br>(1.89) |
| Share of unmarried | 5.03***<br>(1.45) |
| Share of single households | -2.72***<br>(0.83) |
| Share of non-western immigrants | -1.70**<br>(0.70) |
| Share with long-term illness | 1.14<br>(1.04) |
| Mean $PM_{2.5}$ | 6.39***<br>2.42 |
| $\lambda$ | -0.11<br>0.16 |
| $\rho$ | 0.71<br>0.08 |
| R2 | 0.56 |
| AICc | 3736.24 |

Standard Errors in parenthesis, significance levels as: \*\*\* $p < 0.01$ , \*\* $p < 0.05$ , \* $p < 0.1$

#### 6.2.4 Re-estimation using March 31 confirmed hospital admissions per 100,000 inhabitants

Model 5f in table 7 below, re-estimates Model 2 then uses the covariates in the same specification as Model 5 using confirmed hospital admissions from March 31. This is to evaluate whether the relationship with PM_2.5_ is robust to possible measurement error in the confirmed cases. The correlation between confirmed cases per 100,000 inhabitants on March 22 and March 31 hospital admissions is approximately .80, the correlation using March 30 confirmed cases is .88. It is clear from the estimation results that using confirmed admissions instead of cases does not alter the main conclusions. In particular, the parameter estimate for PM_2.5_ remains highly significant.

**Table 7:**
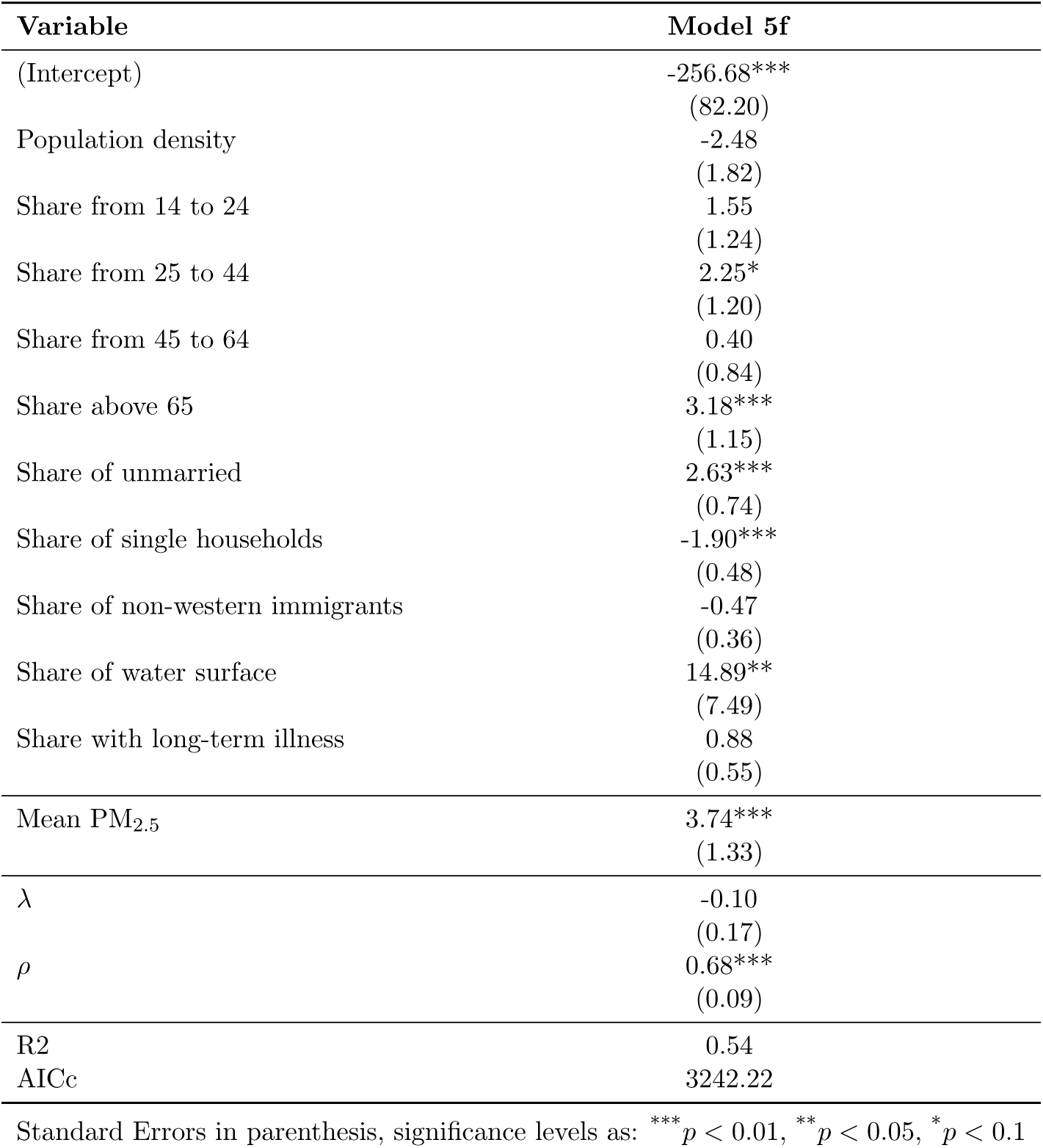
Dependent Variable: Confirmed COVID-19 hospital admissions per 100,000 inhabitants.

#### 6.2.5 Linear analysis of case hospitalization rates

The main results provided evidence for increased COVID-19 incidence in areas where populations are more exposed to air pollution. However, it is possible that the estimated association between PM_2.5_ concentrations and COVID-19 incidence can be attributed to worse respiratory health in polluted areas, which then leads to more severe symptoms and higher case detection. The analysis tried to control for this using health data and the percentage of confirmed cases that resulted in hospitalization as controls. This did not impact the results. A second way to further test this theory is by analyzing the association between PM_2.5_ and the case hospitalization rate because worse respiratory health would lead to more severe COVID-19 disease (Ruan, 2020). The suspect correlation is investigated below using step-wise AIC variable selection keeping PM_2.5_ in the variable set, followed by the full spatial specification. Model 6 uses March 31 confirmed COVID-19 hospital admissions as a percentage of March 22 confirmed COVID-19 cases, model 7 uses March 30 confirmed cases. The analysis finds that age, male gender, and the share of population with overweight are positively associated with increased case hospitalization rates which follows earlier identified risk groups (Ruan, 2020).

**Table 8:**
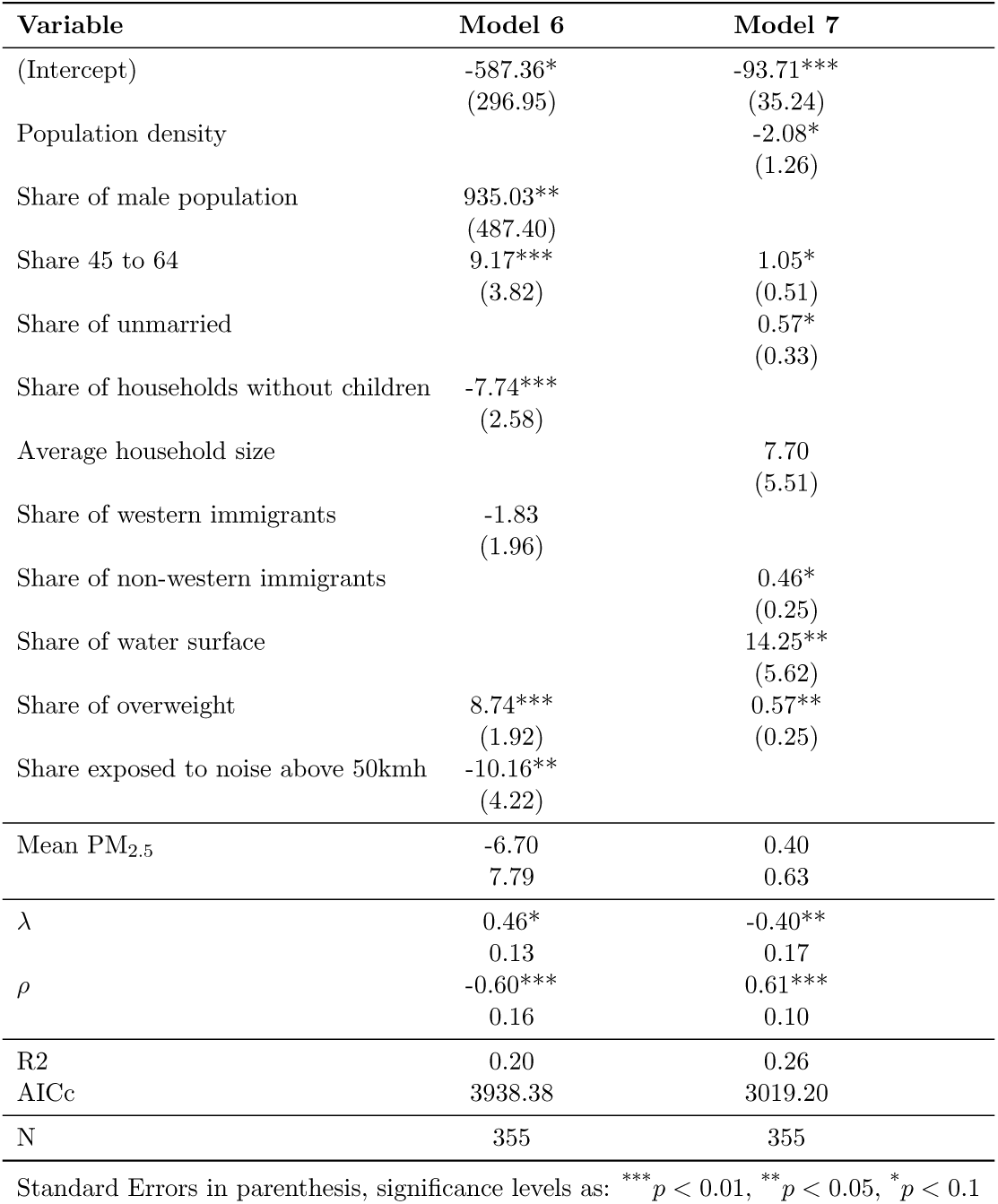
Dependent Variable: Confirmed COVID-19 related hospital admissions as a percentage of COVID-19 cases.

| Variable | Model 6 | Model 7 |
| --- | --- | --- |
| (Intercept) | -587.36*<br>(296.95) | -93.71***<br>(35.24) |
| Population density |  | -2.08*<br>(1.26) |
| Share of male population | 935.03**<br>(487.40) |  |
| Share 45 to 64 | 9.17***<br>(3.82) | 1.05*<br>(0.51) |
| Share of unmarried |  | 0.57*<br>(0.33) |
| Share of households without children | -7.74***<br>(2.58) |  |
| Average household size |  | 7.70<br>(5.51) |
| Share of western immigrants | -1.83<br>(1.96) |  |
| Share of non-western immigrants |  | 0.46*<br>(0.25) |
| Share of water surface |  | 14.25**<br>(5.62) |
| Share of overweight | 8.74***<br>(1.92) | 0.57**<br>(0.25) |
| Share exposed to noise above 50kmh | -10.16**<br>(4.22) |  |
| Mean $PM_{2.5}$ | -6.70<br>7.79 | 0.40<br>0.63 |
| $\lambda$ | 0.46*<br>0.13 | -0.40**<br>0.17 |
| $\rho$ | -0.60***<br>0.16 | 0.61***<br>0.10 |
| R2 | 0.20 | 0.26 |
| AICc | 3938.38 | 3019.20 |
| N | 355 | 355 |

Standard Errors in parenthesis, significance levels as: \*\*\* $p < 0.01$ , \*\* $p < 0.05$ , \* $p < 0.1$

#### 6.2.6 Re-estimation using satellite-derived PM_2.5_

This section evaluates whether the relationship with PM_2.5_ generalizes to measurements from a different source. Table 9 compares the municipality-level data. Both measurements trend in the same direction but the levels according to the RIVM are roughly one-third below those of van Donkelaar.

**Table 9:** Comparison of PM_2.5_ statistics: RIVM 2017 vs van Donkelaar 2016.

| Variable | Mean | Min | q.25 | q.50 | q.75 | Max |
| --- | --- | --- | --- | --- | --- | --- |
| RIVM 2017 | 10.22 | 6.95 | 9.43 | 10.69 | 11.23 | 12.04 |
| van Donkelaar 2016 | 14.60 | 10.15 | 13.44 | 15.01 | 16.01 | 18.05 |

Correlation: 0.70

Model 5g in table 10, estimates step-wise AIC with the new pollution data and uses the selected covariates in the specification of Model 5. Model 5h uses the same controls as Model 5, and Model 5i applies the Yeo-Johnson power transform. The analysis makes use of the confirmed cases from March 22. It is clear from the estimation results that the use of satellite-derived PM_2.5_ results in similar conclusions. The parameter estimate for PM_2.5_ remains significant in the presence of controls and is significant at the highest level when the dependent variable is first normalized.

**Table 10:** Dependent Variable: Confirmed COVID-19 cases per 100,000 inhabitants.

| Variable | Model 5g | Model 5h | Model 5i |
| --- | --- | --- | --- |
| (Intercept) | -127.26*<br>(74.02) | -213.60***<br>(72.86) | -11.17***<br>(2.46) |
| Population density |  | 0.90<br>(1.93) | -0.04<br>(0.07) |
| Share from 25 to 44 |  | -0.50<br>(1.07) | 0.026<br>(0.04) |
| Share above 65 | 2.38***<br>(0.91) | 2.21**<br>(1.06) | 0.10***<br>(0.04) |
| Share of unmarried | 3.50***<br>(1.02) | 4.14***<br>(0.96) | 0.14***<br>(0.03) |
| Share of single households | -1.76***<br>(0.46) | -1.78***<br>(0.50) | -0.07***<br>(0.02) |
| Share of non-western immigrants | -0.53<br>(0.34) | -0.71*<br>(0.41) | -0.02<br>(0.01) |
| Share of water surface | 2.47<br>(8.71) | 2.22<br>(8.75) | -0.79***<br>(0.28) |
| Share of overweight | -1.32<br>(0.91) |  |  |
| Share with obesity | 1.60<br>(1.53) |  |  |
| Share with long-term illness |  | 0.68<br>(0.78) | 0.01<br>(0.02) |
| Mean PM<sub>2.5</sub> | 2.75**<br>(1.28) | 3.13**<br>(1.29) | 0.28***<br>(0.03) |
| $\lambda$ | 0.41*<br>(0.24) | 0.40*<br>(0.24) | 0.10<br>(0.19) |
| $\rho$ | 0.45**<br>(0.22) | 0.46*<br>(0.22) | -0.16<br>(0.19) |
| R<sup>2</sup> | 0.50 | 0.50 | 0.30 |
| AICc | 3265.34 | 3268.53 | 920.81 |
| N | 355 | 355 | 355 |

Standard Errors in parenthesis, significance levels as: \*\*\* $p < 0.01$ , \*\* $p < 0.05$ , \* $p < 0.1$

## Data Availability

The data can be requested from the author.

## Acknowledgments

Thanks to Aart Kraay, Antonio Gasparrini, Urvashi Narain, Sameer Akbar, Michael Toman, Nadia Piffaretti, Phoebe Spencer and participants of the March 9 World Bank “Data Lab Clinic: COVID-19 and Air Quality” for valuable comments and stimulating remarks on an earlier version of the analysis, and Andres Chamorro for managing several of the used data sources. This work has been funded under the World Bank DT4D program. This paper reflects the views of the author, and does not reflect the official views of the World Bank, its Executive Directors, or the countries they represent.

## Footnotes

1. Over the years, numerous studies have related hospitalization numbers, case numbers, and relative risk of respiratory viral infections and influenza-like illnesses to short-term air pollution exposure, mostly at city level, using a variety of data sets and methods. See (Ciencewicki and Jaspers, 2007) for an early review, see (Xu et al., 2013; Liang et al., 2014; Su et al., 2019) on influenza-like illnesses, and (Silva et al., 2014; Huang et al., 2016; Feng et al., 2016; Li et al., 2018) on viral respiratory infections.

2. https://www.rivm.nl/coronavirus-kaart-van-nederland-per-gemeente.

3. https://experience.arcgis.com/experience/478220a4c454480e823b17327b2bf1d4.

4. http://opendatadpc.maps.arcgis.com/apps/opsdashboard/index.html#/b0c68bce2cce478eaac82fe38d4138b1

5. https://github.com/pcm-dpc/COVID-19/tree/master/dati-province

6. https://www.rtve.es/noticias/20200323/mapa-del-coronavirus-espana/2004681.shtml

7. https://www.cbs.nl/nl-nl/nieuws/2018/31/belang-industrie-voor-de-regio

8. https://www.rivm.nl/coronavirus-kaart-van-nederland-per-gemeente

9. For example, early estimates based on Chinese cases indicated that the hospitalization rate of elderly, the most vulnerable population, was only 18.4% (Verity et al., 2020).

10. https://www.cbs.nl/nl-nl/dossier/nederland-regionaal/geografische-data/wijk-en-buurtkaart-2019

11. https://www.rivm.nl/media/smap/langdurigeziekte.html

12. https://www.atlasleefomgeving.nl/kaarten

13. The treatment of spatial autocorrelation and spatial residual correlation took a firm position in quantitative geography after the contributions by Cliff and Ord (1969, 1972). Spatial econometrics as a subfield of econometrics was rapidly developed as a means to analyze sub-country data in regional econometric models (Anselin, 2010). Good introductory books exist, apart from the one referenced in the main text (LeSage and Pace, 2009) is one other. The (Q)MLE is worked out, for example, in (Lee, 2004). The field is still actively developed, with recent advances focusing on time series dynamics and non-linearity (Beenstock and Felsenstein, 2019; Andrée, 2020).

14. The first case was detected on February 27 in Loon op Zand in Brabant, but that same night a case was also confirmed in Amsterdam. Within 4 days, 10 cases had been confirmed in 6 cities across 4 provinces with multiple sources of infection, it took till March 23 for lockdown policies to be announced, giving ample time for spread from multiple points, see https://nos.nl/artikel/2325309-beatrixziekenhuis-gorinchem-gesloten-om-coronavirus-tien-patienten-in-nl.html

15. While the spatial autoregressive models only include the rate of infective subjects in neighboring areas as regressors, the models in fact allow for feedback and spillovers to more distant observations as each area is also a second-order neighbor of itself. See the literature on spatial models cited earlier.

16. The proxy is not perfect due to the low data density. Using instead the percentage of March 30 cases that resulted in March 31 hospitalization leads to only 4 replacements of both types (100/0). Re-estimating Model 5b with this recalculated proxy did not result in measurable change relative to Model 5. Using instead the March 30 confirmed cases as dependent variable in the same regression specification did not find the severity proxy to be significant and found PM_2.5_ to remain significant at the highest level.

